# Detecting sleep in free-living conditions without sleep-diaries: a device-agnostic, wearable heart rate sensing approach

**DOI:** 10.1101/2020.09.05.20188367

**Authors:** Ignacio Perez-Pozuelo, Marius Posa, Dimitris Spathis, Kate Westgate, Nicholas Wareham, Cecilia Mascolo, Søren Brage, Joao Palotti

## Abstract

The rise of multisensor wearable devices offers a unique opportunity for the objective inference of sleep outside laboratories, enabling longitudinal monitoring in large populations. To enhance objectivity and facilitate cross-cohort comparisons, sleep detection algorithms in free-living conditions should rely on personalized but device-agnostic features, which can be applied without laborious human annotations or sleep diaries. We developed and tested a heart rate-based algorithm that captures inter- and intra-individual sleep differences, does not require human input and can be applied in free-living conditions. The algorithm was evaluated across four study cohorts using different research- and consumer-grade devices for over 2,000 nights. Recording periods included both 24-hour free-living and conventional lab-based night-only data. Our method was systematically optimized and evaluated against polysomnography (PSG) and sleep diaries and compared to sleep periods produced by accelerometry-based angular change algorithms. Against sleep diaries, the algorithm yielded a mean squared error (MSE) of 0.04 to 0.06 and a total sleep time deviation of -2.70 (*±*5.74) and 12.80 (*±*3.89) minutes, respectively. When evaluated with PSG lab studies, the MSE ranged between 0.06 and 0.11 yielding a time deviation between -29.07 and -55.04 minutes. Our findings suggest that the heart rate-based algorithm can reliably and objectively infer sleep under longitudinal, free-living conditions, independent of the wearable device used. This represents the first open-source algorithm that can infer sleep using heart rate signals without actigraphy or diary annotations.

## 1. Introduction

Human sleep is a reversible physiological state that is essential for health and performance [1]. The functions of sleep are not fully understood, but its influence on energy homeostasis, brain function, cognitive performance and behaviour, as well as interactions with the immune system, promotion of healing and consequences for numerous health conditions have been studied extensively [2–10]. As such, sleep lies at the cross-roads of multiple research programs in both life sciences and public health. This makes the objective monitoring of sleep crucial for understanding human health and well-being. Apart from outright sleep disorders, changes to sleep patterns impact the quality of life and the history of common disease, whether cardiovascular, metabolic or neurodegenerative. The gold-standard method to quantify sleep quantity and quality is polysomnography (PSG). PSG requires signals from multiple sensors, as well as expert technician and clinician input and is thus limited to the laboratory setting. This is resource-intensive and limits large-scale and long-term population studies. Furthermore, the sleep recorded in the unfamiliar laboratory environment might not reflect the patients’ typical sleep pattern [11].

Actigraphy is a well-established and widely-used alternative to PSG. It originates in early telemetric measurements of motor activity in the 1970s which were used to assess sleep quality [12]. Since then, a vast number of studies have assessed the use of actigraphy for sleep monitoring against PSG [13–15]. The advantage over PSG is that actigraphy, as well as its modern counterpart, accelerometry, require sensors that can be integrated into more affordable wrist-worn devices [13, 16]. At present, ambulatory and longitudinal monitoring of healthy sleepers through actigraphy is approved by both the FDA and recommended by the American Academy of Sleep Medicine (AASM) [14].

Over the past 30 years, a number of actigraphy-based algorithms have been developed to detect night-time sleep and wake periods. A few have proved to have strong validity and reliability against PSG [17–22]. These algorithms have been widely adopted and were recently bench-marked against both each other and newer machine learning and deep learning methods, which highlighted the strengths and limitations of each method [23, 24]. For example, across multiple studies it was shown that actigraphy struggled to classify wake events during the sleep period, yielding poor specificity compared to PSG [16, 21, 23, 25, 26]. Additionally, the actigraphy algorithms have only been optimised to detect sleep during the night, as study participants typically only spend a night or two in the laboratory. Using them with data recorded over the entire day is unreliable, even with sleep annotations provided by PSG technicians or the subjects themselves [27, 28]. This need for manually restricting sleep search windows is also present in most proprietary algorithms used by commercial wearables. In addition, actigraphy devices do not provide real-time feedback about the user’s sleep, which makes longitudinal monitoring cumbersome. The result is that sleep patterns that are different from the monophasic night sleep recorded in laboratories, whether due to cultural differences or shift work have been understudied.

Novel wearable devices have started to combine actigraphy/accelerometry and heart rate signals, via photoplethysmography (PPG). These multimodal devices rely on recent advances in microelectromechanical systems (MEMS) along with improvements in cost, battery capacity, and increased memory, which allows for a higher sensor sampling rate. The widespread adoption of these devices for both research and commercial use promise more robust inferences about the user’s sleep and awake periods. To this end, large investments have been made both by technology companies looking to offer personal health monitoring and through research grants for programs such as “All of US” [29]. This increased attention has made the need to validate data collected by such wearables against gold-standard PSG imperative [30], especially as recent studies have claimed not only to predict the presence of sleep but also to label which sleep stage the user is in at any time [31, 32]. Furthermore, as many studies aim to look into cardiovascular disease, incorporating heart rate (HR) and heart rate variability (HRV) attempts to bring into sleep algorithms predictors that are directly linked with cardiovascular fitness [24, 33, 34].

Whilst these approaches are valuable, they have limited applicability in other large, free-living cohort studies for three main reasons. First, they rely on machine learning methods derived specifically for those datasets. Domain adaptation is needed before applying them to a different population or even the same population using a different device. Second, just like all the previous algorithms, they are only derived from night-time data, limiting their generalizability to less regular sleep patterns [35]. Finally, these approaches still rely on self-reported input about habitual sleep times, either through questionnaires or sleep diaries (time of going to sleep and waking up). While questionnaires are prone to recall bias and only provide data about the previous few weeks, sleep diaries are tedious to keep for more than a few days [36, 37]. Even when sleep diaries are available, it can take in excess of 6 recorded days to achieve an agreement with objective labels, even amongst those with more regular sleep patterns [38]. This time is close to the usual time study participants are asked to wear the multimodal devices, which is mostly contingent on device battery life. An approach which can dispense with laborious annotations can make inferences more reliable, encouraging wider adoption by long-term users. This can, in turn, provide longitudinal data from devices that are familiar to participants in free-living conditions. These benefits can extend the scope of existing trials in terms of recruitment, data collection period and cost, while improving the inferences users can obtain about their sleep.

Our approach leverages heart rate data available from most commercial and research-grade wearable devices to develop a sleep detection algorithm. The algorithm is device-agnostic and provides the quantitative sleep and awakening inferences offered by previous methods. We validated the algorithm, including the lack of reliance on sleep annotations, against multiple datasets where heart rate data was recorded together with multiple PSG-grade sensors or actigraphy. As our algorithm does not require training before deployment on each individual device, as do machine learning methods, it can run on the device itself without the need for cloud computing. This enhances the user’s privacy, which is paramount for health data. The approach was evaluated in four separate settings that used different wearable devices. First, the algorithm was developed in a large population (n=193) with about 8 nights of recording accompanied by detailed sleep diaries. This cohort wore a combined heart rate and movement sensor, in addition to a set of 3 accelerometers on both wrists and hip. This gave us both the diary annotations and the chance to compare our heart rate-based approach with previous methods based on accelerometer angle changes across all the usual anatomical locations used for such devices. As data from multiple consecutive days and nights was available, this dataset facilitated testing for inter- and intra-individual variability (i.e., sleep statistics across the entire cohort or across each participant’s sleep windows). We then assess our method in a larger, more diverse, open-source dataset (n=1,743), as well as a smaller cohort (n=31) that used a commercial-grade device (Apple Watch) during PSG. Finally, the performance in free-living conditions was also tested against detailed sleep diaries in an accelerometer/heart rate-sensor wearing cohort (n=22).

## 2. Results

### 2.1. Participant characteristics

The aim of our study was to explore the feasibility and effectiveness of a device-agnostic, heart-rate based algorithm to classify sleep periods in the absence of sleep diaries or annotations. To this end, we selected one of the cohorts (BBVS) as benchmark for optimal parameter search, as it contains data for the entire day. However, much of the annotated data in other cohorts (e.g., MESA) from academic studies only covers the night period. We matched this structural difference between datasets by analysing the BBVS cohort separately for the entire day and for the night-time period only. This is to show that differences in the best parameters stem from the fact that night-time only data often does not come from free-living conditions and requires separate parameter inference available via the heart-rate algorithm’s built-in capabilities. The optimal parameters from the 24-hour and night-time BBVS analysis were then applied to the other 3 cohorts. Cohort and evaluation and validation set characteristics are summarized in Table 1.

**Table 1:** Summary of data set demographics.

| Feature | Biobank Validation Study (BBVS) | Multi-Ethnic Study of Atherosclerosis (MESA) | PhysioNet Apple Watch | Multilevel Monitoring of Activity and Sleep in Healthy people (MMASH) |
| --- | --- | --- | --- | --- |
| *Number of participants* | 193 | 2230 | 31 | 22 |
| *Age (years, mean(std))* | 54.13 (6.95) | 68.65 (8.91) | 29.42 (8.52) | 26.05 (7.12) |
| *Percent Male* | 54.40 | 46.28 | 32.26 | 100 |
| *BMI (mean(std))* | 26.21 (3.21) | Not available | Not recorded | 23.12 (3.09) |

### 2.2. Evaluation of the algorithm in the BBVS

The results of the hyper-parameter search on the BBVS dataset are summarized in Table 2 and histograms detailing the results are shown in Figures S2 and S3. While we envision that our algorithm’s larger contribution is when using it on full-day, we also experiment with data only for the night period. The hyper-parameters picked by the grid-search method are used in all the other experiments in this report to show our approach’s generalizability.

The set of hyper-parameters vary depending on the scenario that they are used. For the whole day experiment, the best results were achieved when using an HR quantile threshold of 0.325, which resembles the fact that one-third of the day is spend sleeping when the expected HR values are lower. For the night-only experiment, both the HR quantile and the gap-merging threshold are much higher, indicating that the best results were achieved when the algorithm prioritized a monolithic sleep block and aimed only to ignore the activity before sleep onset and after sleep offset.

**Table 2:** Optimal hyper parameters extracted from a grid search on the BBVS dataset for both full-day and night-only data. These parameters are used accordingly on the other three datasets studied in this work.

| Scenario | HR Quantile Threshold (Q) | Minimum Length (L) | Gap Merging Threshold (G) |
| --- | --- | --- | --- |
| Full Day | 0.325 | 20 | 90 |
| Night Only | 0.800 | 20 | 420 |

The results of the evaluation on the BBVS study are summarized in Table 3. Our HR algorithm optimized on the full day yielded an MSE of 0.06 and estimated a TST on average 9.84 minutes longer compared to sleep diaries, when looking at the entire 24-hour recording period. We compared this result with the angular change approach shown in Table 4. The best performing wrist-worn device (non-dominant wrist) had an overestimation of 192 minutes compared to the sleep diaries. The results across all three accelerometers for this approach were comparable as summarized in Table 4, each yielding an MSE of 0.17.

**Table 3:** Results of applying the HR algorithm on the BBVS dataset for both full-day and night-only data. Comparisons are made against sleep diaries. BBVS TST for diaries mean *±* 95% CI = 7.739 *±* 0.073 hours (464.34 *±* 4.38 minutes).

| Sleep parameter | Metric | HR algorithm (Full day)<br>(mean ± 95% CI) | HR algorithm (Night only)<br>Value (mean ± 95% CI) | p-value |
| --- | --- | --- | --- | --- |
| Total sleep time | Time difference (minutes) | $-2.70 \pm 5.74$ | $12.80 \pm 3.89$ | <0.00 |
| | MSE | $0.06 \pm 0.00$ | $0.04 \pm 0.00$ | <0.00 |
| | Cohen's kappa | $0.86 \pm 0.00$ | $0.90 \pm 0.00$ | <0.00 |
| Sleep onset | Time difference (minutes) | $-0.49 \pm 5.67$ | $-4.59 \pm 3.27$ | 0.158 |
| Sleep offset (Wake Up) | Time difference (minutes) | $-3.19 \pm 4.80$ | $8.20 \pm 2.88$ | <0.00 |

**Table 4:** Comparison of angle algorithm performance for the BBVS dataset by the limb on which the device was worn. All participants wore devices on their dominant (dw) and non-dominant (ndw) wrist as well as on their thigh. The best performance metrics were obtained for the non-dominant wrist device, but thigh wearables gave the least time differences overall in terms of total sleep time (TST), sleep onset and offset. BBVS TST for diaries mean *±* 95% CI = 7.739 *±* 0.073 hours (464.34 *±* 4.38 minutes).

| Sleep parameter | Metric | Angle change algo. (ndw)<br>Value (mean ± 95% CI) | Angle change algo. (dw)<br>Value (mean ± 95% CI) | p-value<br>ndw-dw | Angle change algo. (Thigh)<br>Value (mean ± 95% CI) | p-value<br>ndw-thigh |
| --- | --- | --- | --- | --- | --- | --- |
| Total sleep time | Time difference (min.) | $222.64 \pm 7.78$ | $218.96 \pm 7.92$ | 0.271 | $214.90 \pm 8.08$ | 0.048 |
| | MSE | $0.16 \pm 0.00$ | $0.16 \pm 0.00$ | 0.052 | $0.16 \pm 0.00$ | 0.291 |
| | Cohen's kappa | $0.58 \pm 0.01$ | $0.59 \pm 0.01$ | 0.027 | $0.58 \pm 0.01$ | 0.650 |
| Sleep onset | Time difference (min.) | $-100.99 \pm 6.63$ | $-96.00 \pm 7.01$ | 0.167 | $-96.07 \pm 7.85$ | 0.250 |
| Sleep offset (Wake Up) | Time difference (min.) | $121.65 \pm 7.05$ | $122.96 \pm 7.43$ | 0.727 | $118.82 \pm 8.11$ | 0.501 |

Our HR model estimated sleep onset on average 8.84 minutes later than sleep diary while the angular change approach on the non-dominant wrist resulted in an average overestimation of 87.88 minutes. For diary-based sleep offset, our HR algorithm the estimation was on average 1.00 minute earlier, while for the angular change approach that estimation was 104.33 minutes earlier for the non-dominant wrist. Further, modified Bland-Altman plots for the HR and angle approaches against sleep diary for the BBVS cohort are presented in Figure 1. Finally, Figure 2 showcases an example BBVS participant to whom the HR and angle change sleep algorithms were applied.

**Figure 1:**
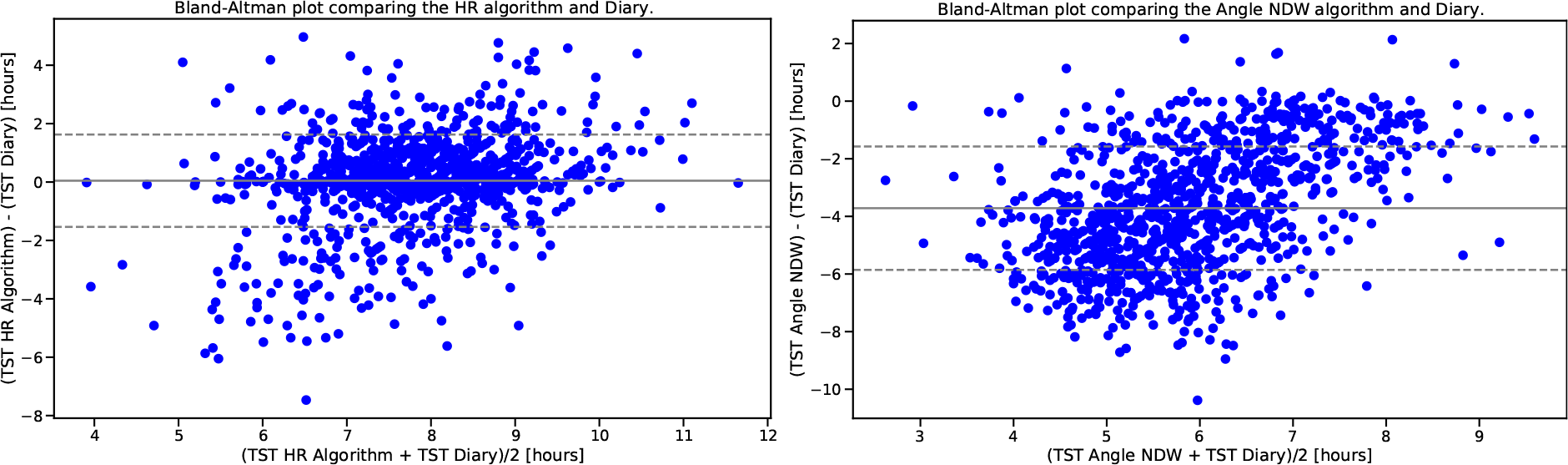
Modified Bland-Altman plot for BBVS. Modified Bland-Altman plot on the left shows the TST differences (delta) between the full-day HR algorithm and diary in the Y-axis and the X-axis shows the TST average for every participant. The figure to the right shows the same comparison for the angle algorithm and diaries in BBVS. Dashed lines represent limits of agreement (LoA) which are defined as the mean difference *±* 1.96 SD of differences. TST: total sleep time.

**Figure 2:**
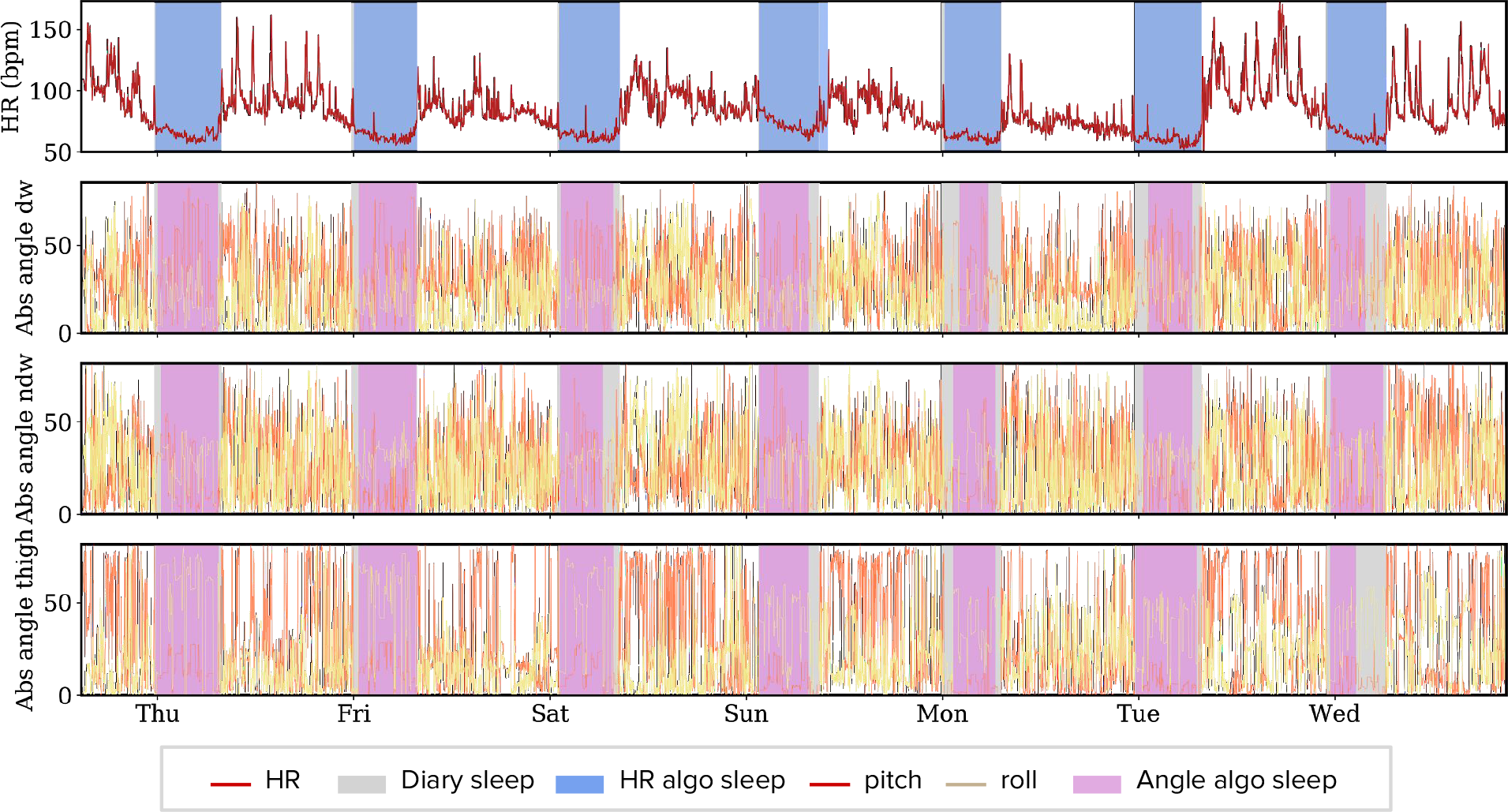
Example participant (chosen at random), showcasing estimated sleep through the heart rate sleep window algorithm, sleep diary sleep onset and offset and angle changes for both wrists and the thigh accelerometers. The algorithm picks up subtle sleep regularity differences at a participant level. This approach overlaps more closely to the sleep diary than any of the accelerometer-based approaches. Notice that, for the angle change approach, the algorithm is more effective on the non-dominant wrist accelerometer than on the dominant wrist or thigh accelerometer for most nights. TST: total sleep time.

### 2.3. Evaluation and fine-tuning of the algorithm in the MESA study

To validate the approach used on the BBVS cohort, we used the MESA study dataset, as both sleep diary and PSG data was available. In addition, the MESA cohort also contains participants formally diagnosed with sleep disorders. The results are detailed in Table 5. Thus, our HR-based algorithm and annotations from sleep diaries and PSG were tested against one another, resulting in the modified Bland-Altman plots in Figure 3.

**Figure 3:**
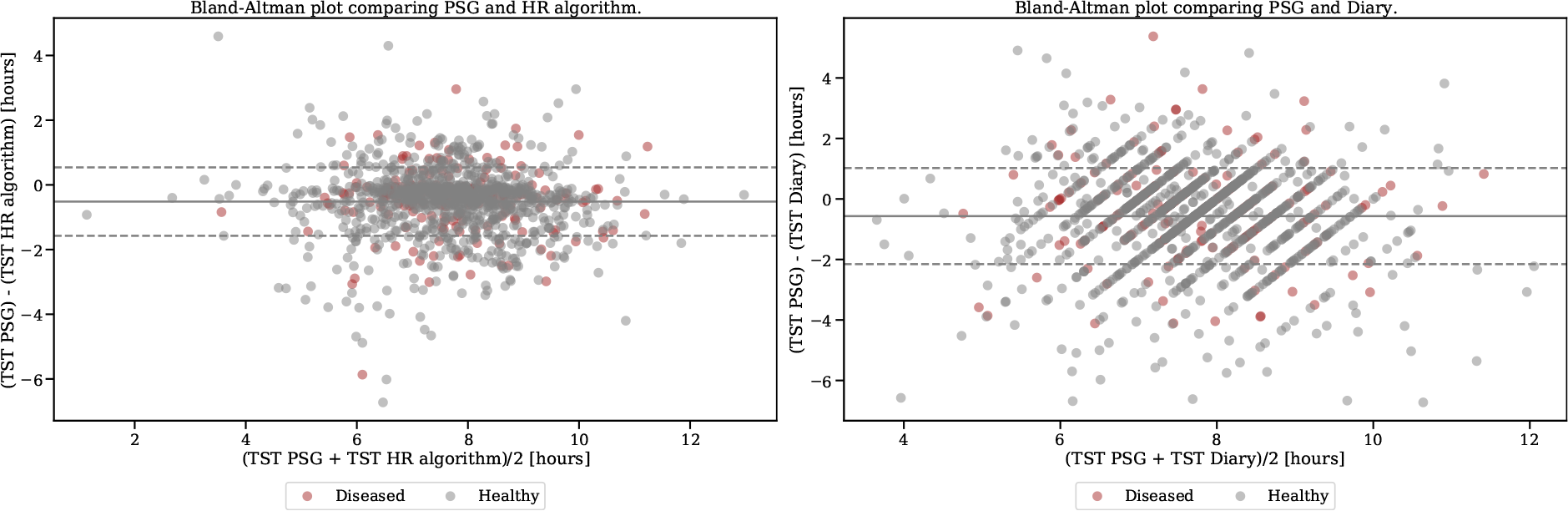
Modified Bland-Altman plot for MESA. Modified Bland-Altman plot on the left shows the TST differences (delta) between the HR algorithm and PSG in the Y-axis and the X-axis shows the TST average for every participant. The figure to the right shows the same comparison for the sleep diaries and PSG in MESA. Further, healthy participants are color coded in blue for both plots and participants that were diagnosed with sleep disorders are shown in orange.

**Figure 4:**
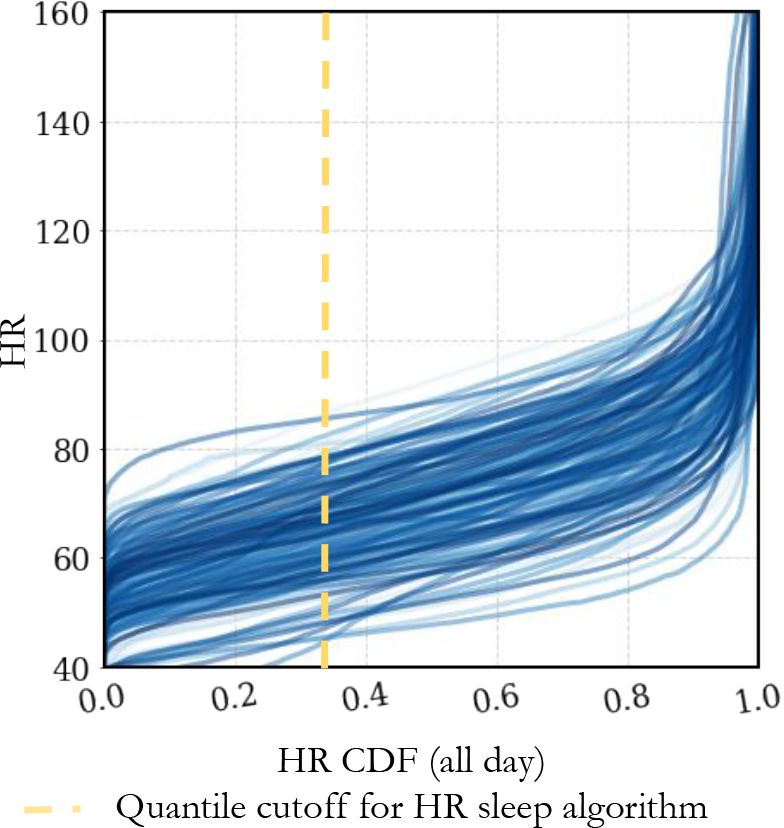
Cumulative distribution function for BBVS heart rates. The figure shows the HR ECDF for the full-day across all participants and all days, where the yellow dotted line shows the 0.35 HR quantile cutoff. Each individual line represents one participant for one day of recording.

**Table 5:** Results for the MESA dataset. Both the HR algorithm and sleep diaries are evaluated against PSG. Results are also shown for the subset of healthy participants and participants with sleep disorders. MESA TST for PSG mean *±* 95% CI = 7.433 *±* 0.079 hours (445.95 *±* 4.71 minutes). N=1,154

| Sleep parameter | Metric | HR algorithm | Sleep Diary | p-value |
| --- | --- | --- | --- | --- |
| | | Value (mean ± 95% CI) | Value (mean ± 95% CI) | |
| Total sleep time | Time difference (min.) | $-55.04 \pm 3.75$ | $-34.04 \pm 5.50$ | $<0.00$ |
| | MSE | $0.11 \pm 0.01$ | $0.13 \pm 0.01$ | $<0.00$ |
| | Cohen’s kappa | $0.59 \pm 0.02$ | $0.62 \pm 0.01$ | 0.01 |
| Sleep onset | Time difference (min.) | $39.72 \pm 3.01$ | $6.25 \pm 3.30$ | $<0.00$ |
| Sleep offset (Wake Up) | Time difference (min.) | $-15.32 \pm 2.52$ | $-27.79 \pm 4.86$ | $<0.00$ |
| Healthy participants (N = 965) |  |  |  |  |
| Total Sleep Time | Time difference (min.) | $-56.12 \pm 4.32$ | $-36.05 \pm 6.05$ | $<0.00$ |
| | MSE | $0.11 \pm 0.01$ | $0.13 \pm 0.01$ | $<0.00$ |
| | Cohen’s kappa | $0.59 \pm 0.02$ | $0.62 \pm 0.02$ | 0.013 |
| Sleep onset | Time difference (min.) | $40.36 \pm 3.30$ | $6.12 \pm 3.64$ | $<0.00$ |
| Sleep offset (Wake Up) | Time difference (min.) | $-15.76 \pm 2.78$ | $-29.93 \pm 5.34$ | $<0.00$ |
| Participants with sleep disorders (N = 189) |  |  |  |  |
| Total Sleep Time | Time difference (min.) | $-49.51 \pm 9.47$ | $-23.75 \pm 13.05$ | $<0.00$ |
| | MSE | $0.11 \pm 0.01$ | $0.13 \pm 0.01$ | 0.071 |
| | Cohen’s kappa | $0.58 \pm 0.04$ | $0.60 \pm 0.04$ | 0.448 |
| Sleep onset | Time difference (min.) | $36.45 \pm 7.41$ | $6.92 \pm 7.82$ | $<0.00$ |
| Sleep offset (Wake Up) | Time difference (min.) | $-13.07 \pm 5.96$ | $-16.84 \pm 11.66$ | 0.540 |

Results from the MESA cohort confirmed that the HR-based algorithm was non-inferior to human-annotated sleep. For both healthy sleepers and participants with sleep disorders, the average MSE for the PSG sleep-wake labels was 0.11 (versus 0.13 for sleep diaries). The superior performance of using HR was also reflected in a better Cohen’s kappa for all three analyses. Both approaches underestimated the total sleep time compared to PSG-derived labels by -55.04 and -34.04 minutes, respectively for the HR algorithm and sleep diaries, a difference that was statistically significant. Interestingly, our algorithm was better at inferring sleep offset (-15 minutes compared to PSG labels, versus about -30 minutes for sleep diaries), but worse at detecting sleep onset (40 minutes, versus 6 minutes for the sleep diary). As the MESA study only recorded night-time data, the HR quantile optimal for the 24-hour BBVS data was not suitable. Instead, we benchmarked the MESA data against the best HR quantile for a night-only window of the BBVS cohort, which was 0.80. This lead to an MSE of 0.11 between the HR algorithm and PSG sleep labels.

### 2.4. Validation of the algorithm in the PhysioNet Apple Watch Polysomnography study

Our algorithm was applied to data obtained from a commercial, readily available wrist-worn wearable and evaluated against gold-standard measures of sleep obtained with PSG. In this cohort, we evaluated both the HR algorithm and the angle change approach given the presence of triaxial accelerometry. Using the optimal parameters derived from the night-only BBVS data, the HR algorithm resulted in an MSE of 0.07 while the wrist-based angular change approach yielded an MSE of 0.12. Total sleep time deviation was of -29.07 minutes for the HR approach and 44.39 for the angle change approach. Sleep onset time deviation was of 20.73 minutes for the HR approach and of -21.77 for the angle change approach, while the difference was of -8.34 and 22.61 for sleep offset. However, Cohen’s kappa was slightly lower for the HR approach (0.59) than for the angle change algorithm (0.71). These results are summarized in Table 6.

**Table 6:** Results for the PhysioNet Apple Watch dataset. The table presents results for both the HR and angle change algorithm for total sleep time, sleep onset and sleep offset in the PhysioNet Apple Watch dataset. PhysioNet Apple Watch TST for PSG mean *±* 95% CI = 7.165 *±* 0.544 (429.89 *±* 32.65 minutes). ndw: Non-dominant Wrist. N=22

| Sleep parameter | Metric | HR Algorithm | Angle change algorithm (ndw) | p-value<br>(n = 22) |
| --- | --- | --- | --- | --- |
| | | Value (mean ± 95% CI) | Value (mean ± 95% CI) | |
| Total sleep time | Time difference (minutes) | $-29.07 \pm 13.38$ | $44.39 \pm 40.01$ | 0.001 |
| | MSE | $0.07 \pm 0.03$ | $0.12 \pm 0.08$ | 0.277 |
| | Cohen’s kappa | $0.59 \pm 0.12$ | $0.71 \pm 0.13$ | 0.234 |
| Sleep onset | Time difference (minutes) | $20.73 \pm 5.45$ | $-21.77 \pm 29.77$ | 0.008 |
| Sleep offset (Wake Up) | Time difference (minutes) | $-8.34 \pm 11.98$ | $22.61 \pm 31.01$ | 0.056 |

### 2.5. Validation of the algorithm in the MMASH study

Our final set of evaluations took place in the MMASH cohort, which included both HR and triaxial accelerometer data recorded continuously for full-day periods. We evaluated both the proposed HR approach and the angle change approach against detailed sleep diaries. Starting from the 24-hour BBVS data-derived best parameters, we obtained an MSE of 0.11 and total sleep time difference of 17.64 minutes, with a Cohen’s kappa of 0.75 for the HR approach. As additional validation, we performed the same analysis on MMASH data from the night period only, using the night-only BBVS best parameters, with a similar MSE result of 0.09 against sleep diaries. On the other hand, the angle change approach resulted in an MSE of 0.10 and Cohen’s kappa of 0.78, but the total time deviation was substantially worse, yielding a total sleep time difference of -55.86 minutes. Full results for the MMASH cohort are presented in Table 7. Similar to BBVS, results of the optimal parameter search for the MMASH cohort can be found in the Supplementary Material Figure S4.

**Table 7:** Results for the MMASH dataset. The table presents results for both versions of the HR algorithm and compares them to the angle change algorithm for total sleep time, sleep onset and sleep offset in the MMASH dataset. MMASH TST for diaries mean *±* 95% CI = 6.200 *±* 0.622 hours (371.98 *±* 37.33 minutes). ndw: Non-dominant Wrist. N = 21.

| Sleep param. | Metric | HR Algo. Full Day - HRD | Angle change Algo. (ndw) | p-value<br>(ndw - HRD) | HR Algo. Only Night - HRN | p-value<br>(ndw - HRN) |
| --- | --- | --- | --- | --- | --- | --- |
| | | Value (mean ± 95% CI) | Value (mean ± 95% CI) | | Value (mean ± 95% CI) | |
| Total sleep time | Time difference (min) | 17.64 ± 47.78 | -55.86 ± 42.67 | 0.009 | -34.36 ± 35.24 | 0.366 |
| | MSE | 0.11 ± 0.04 | 0.10 ± 0.04 | 0.487 | 0.09 ± 0.03 | 0.742 |
| | Cohen's kappa | 0.75 ± 0.10 | 0.78 ± 0.09 | 0.465 | 0.80 ± 0.06 | 0.692 |
| Sleep onset | Time difference (min) | -39.14 ± 44.60 | 9.55 ± 34.87 | 0.127 | 36.00 ± 24.85 | 0.204 |
| Sleep offset | Time difference (min) | -21.50 ± 33.85 | -46.31 ± 31.64 | 0.080 | 1.64 ± 33.09 | <0.00 |

## 3. Discussion

Objective and unobtrusive measurement of sleep in large, free-living populations at scale will help facilitate epidemiological investigations powered to explore the relationships between sleep, physical behaviours and disease. Concurrently, the rapid growth and adoption of commercial grade wearable devices offers a unique opportunity for the objective monitoring of sleep at scale. However, most commercial devices use algorithms that are not open-source or do not report thorough validation against gold-standard measures. Similarly, conventional algorithms tend to rely on device specific metrics, such as counts, requiring extensive adaptation for each device and cohort tested, as well as a predefined search window through expert annotations or sleep diaries. This often renders evaluation across devices and without sleep diaries futile.

Here we introduced a device agnostic algorithm that exploits the HR sensing capabilities present in most modern wearable devices. We presented an algorithm based on a personalized HR feature that allows detection of sleeping windows under free-living conditions. The proposed method relies on the well-established changes in HR that occur when individuals transition from wake to sleep [39]. Hence, our approach is likely able to infer sleep on individuals regardless of fitness level or illness and could be used amongst shift workers who exhibit sleep episodes outside of the night period (as explored in Figure 6). These qualities may be particularly relevant when evaluating sleep in populations with fragmented sleep, in countries where sleep timing changes due to seasonality or where cross-cultural sleep differences are observed [40]. The value of this approach lies in the fact that it is device-agnostic, does not require sleep diary or questionnaire data and adapts to inter- and intra-individual (day-to-day) variability, allowing for accurate and reliable sleep window labeling.

**Figure 5:**
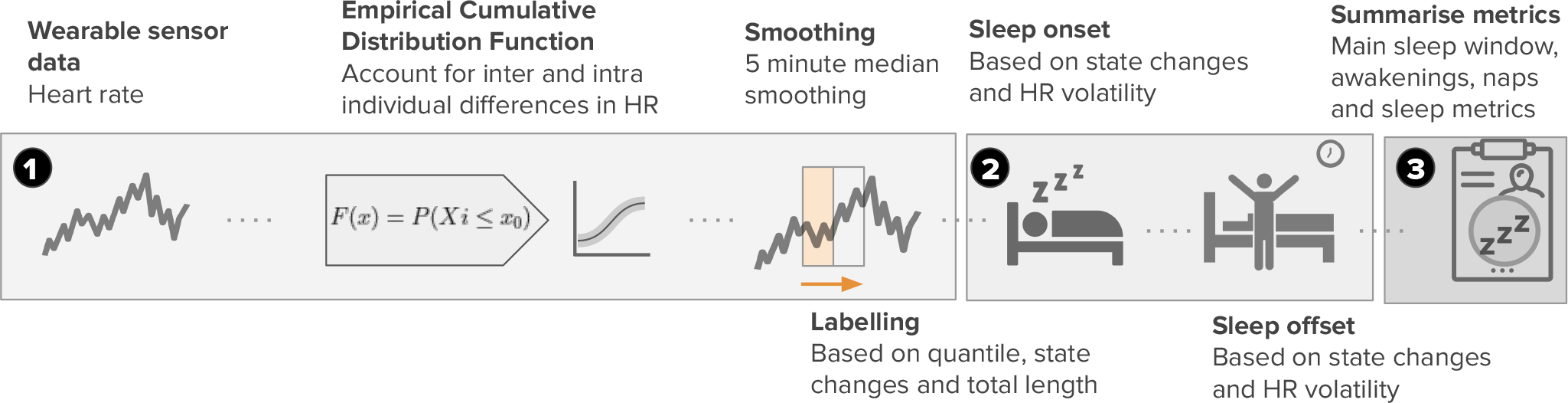
Heart rate sleep algorithm description. The approach can be broken down into three distinct steps. The first step, involves obtaining the wearable sensor HR data, pre-processing that data and setting initial sleep blocks through ECDF quantile thresholds *Q*. Blocks longer than *L* minutes are kept and merged with other blocks if their gap is smaller than *G* minutes. We extract the limits of the resulting blocks as sleep candidate for sleep onset and offset. Next, rolling heart rate volatility is used to refine these candidate times by finding nearby periods where this volatility is high. Finally, nap and awakenings are labeled, the former coming from the candidate sleep blocks not included in the largest sleep window, while the latter are short periods (*<*60 minutes) within the sleep window when the heart rate exceeds the daytime threshold. A detailed description of this algorithm and parameters used can be found in the methods section.

**Figure 6:**
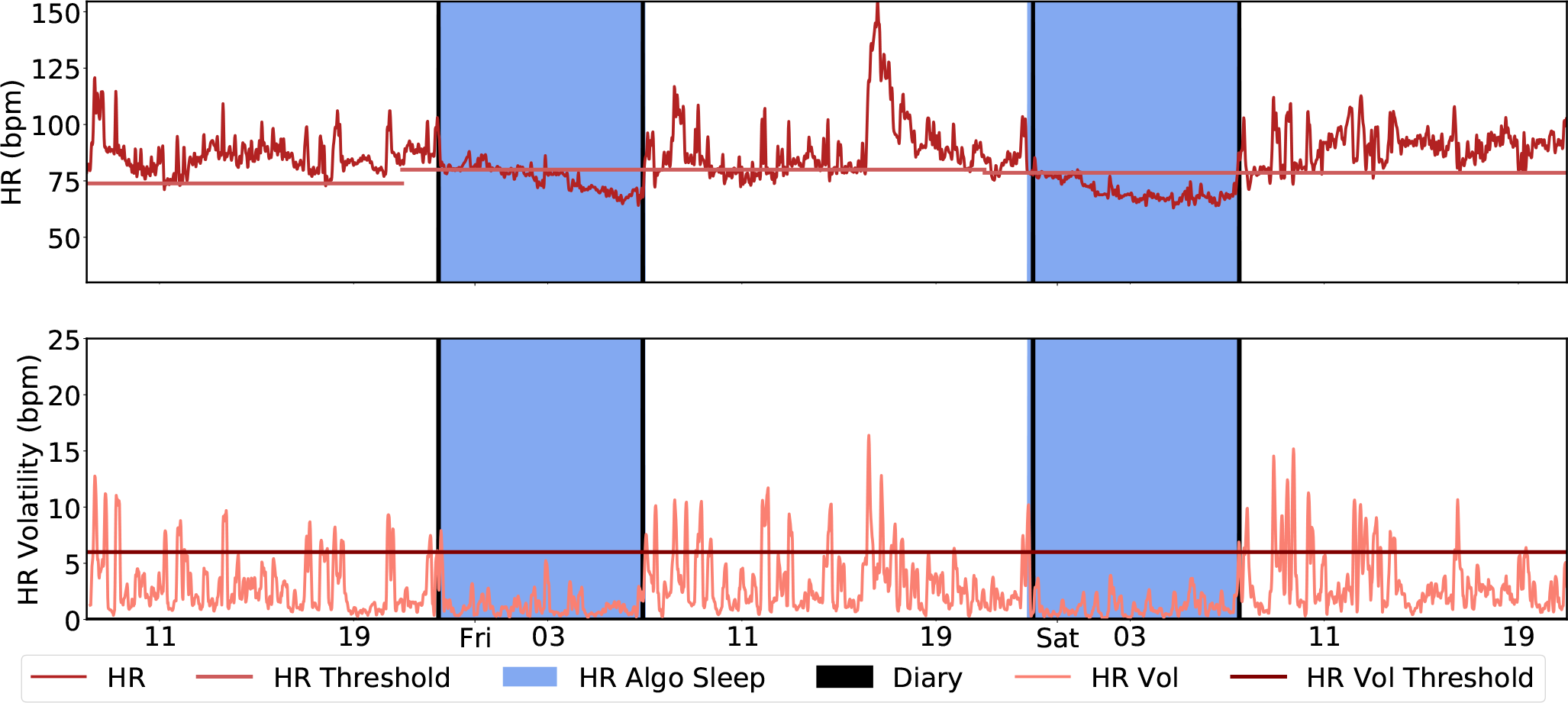
Heart rate sleep algorithm in action for a participant chosen at random. The first step involves setting initial sleep blocks through ECDF quantile thresholds (in this experiment, *Q* = .35). Blocks longer than *L* = 40 are kept and merged if the gap between blocks is smaller than *G* = 60 minutes. We extract the limits of the resulting blocks as candidate state changes. The bottom panel highlights the use of rolling heart rate volatility to refine these candidate times by finding nearby periods where this volatility is high. The resulting candidate times designate each day’s main sleep window.

One important caveat to note is that the algorithm is also capable of inferring naps and fragmented sleep episodes. Naps are denoted by sleep periods outside of the buffer 180-window either side of the main sleep window that are less than 90 minutes in length. To account for polyphasic sleepers and insomniacs, sleep episodes that are outside of that window and longer than 90 minutes are also considered sleep blocks.

We evaluated our HR-based algorithm in four cohorts: BBVS, MMASH, PhysioNet and MESA. Both BBVS and MMASH include free-living HR, movement and sleep diary data for multiple days. By contrast, PhysioNet and MESA provide lab-based HR data and gold-standard PSG. Our aim was to evaluate the algorithm’s performance in free-living conditions in the first cohort and compare it to existing measures that could be leveraged in these cohorts, whilst using the other cohorts to evaluate its validity against gold-standard measures. Further, through this process, we aimed to identify the range of parameters (*Q, L, G*) that produce the best results in free-living conditions, allowing for application and deployment in the absence of any ground truth.

For the first evaluation in the BBVS study, we found that the proposed method performed strongly in free-living conditions, with an average time deviation for total sleep time compared to non-habitual sleep diaries of 2.70 minutes. In this study, we performed optimal parameter search using both full-day measures of HR as well as night-only measures to analyze how the availability of sensor data or the design of the experiment affect the choice of best parameters. The parameter search for the optimal MSE was performed based on quantile, window merge and window length values and are presented in Figure S2. We found that the optimal full-day parameters for this cohort were 0.325 for the quantile (*Q*) and 20 minutes for the window length (*L*) and time merge block of 90 minutes (*G*). This resulting optimal quantile, 0.325, also makes intuitive sense as it represents about 8h, which is around the expected time spent sleeping for most individuals in a day. For the night-only data, the best parameters were 0.80 for the quantile, 20 minutes for the window length and a time merge block of 420. The higher quantile for the night period makes intuitive sense as a lower percentage of the total time would have been spent in active behaviours and participants would have been more likely to be sedentary and supine later in the day.

The MSEs against sleep diaries were comparable for night-only and full-day data (0.04 vs. 0.06), which shows that a window-agnostic analysis does not lead to a significant loss of performance. This flexibility allows discovery of non-standard sleep patterns, like biphasic sleep or daytime sleep in shift workers, which would otherwise be ignored by night-only approaches.

The algorithm performed better at detecting self-reported sleep offset (wake up), than sleep onset, yielding a time difference between 1.64 (MMASH) and -15.32 (MESA) and between -0.49 (BBVS) and 39.72 (MESA) minutes, respectively. These results may be affected by two factors. Firtst, due to the fact that the sleep diaries for validation of sleep onset and offset, while more detailed than traditional sleep diaries, rely on self-report and may not be wholly accurate. While sleep offset is relatively straightforward to annotate as most people wake up with alarm clocks, the exact time of sleep onset cannot be recorded, and is prone to measurement bias, if attempted at the time, or recall bias, if filled in the next day. Thus, the quality of self-reported sleep may vary based on the sleep onset latency of each participant for each given night. Second, the considerable differences in the MESA dataset are likely due to the fact that the experiment starts when the participant is already in bed and lying down, yielding limited variance on the HR signal as opposed to the full-day approaches used in the other datasets. Nevertheless, the performance of the method across a diverse population and multiple nights of recording showcases its potential for free-living applications.

Finally, in the BBVS cohort, we evaluated the performance of an angle change-based algorithm inspired by previous work [27, 41] leveraging the multiple accelerometers available to evaluate angle-based postural changes. We found that this approach is valuable, but the results were more modest than those of our proposed method, yielding a total sleep time MSE of 0.16 and a time deviation of 222.64 minutes for the non-dominant wrist device. We also found that using the combined pitch and roll approach versus only the z-angle did not significantly alter the results. In sum, while valuable, the angle change approach performed significantly worse than our HR-based algorithm in the BBVS cohort. These results suggest that when HR is available, it should be used in preference, but triaxial accelerometry is a valuable second option in the absence of HR.

The algorithm was also evaluated in the MESA cohort, a large, diverse population where gold-standard PSG sleep measures through PSG were available, alongside self-reported sleep (through sleep diaries). When analyzing the MESA data, we started from the BBVS night-only optimized parameters, with additional segmentation into healthy and sleep disorder-diagnosed population subsets. This HR algorithm analysis yielded the results reported in Table 5. In MESA, the deviation of total sleep time versus gold-standard measures of sleep was -55.04 minutes and MSE of 0.11 for the full population, whereas the same comparison between PSG and sleep diaries yielded a total sleep time deviation of -34.04 minutes and MSE of 0.13. This shows that our HR-based method can reliably and objectively monitor sleep in the absence of PSG and performs better than sleep diaries. It is also worth noting that the HR approach was better at detecting PSG measured sleep offset (wake up) with a time difference versus PSG of -15.32 minutes compared to the -27.79 of the diaries. These results further highlight that our algorithm can be used in the absence of sleep diaries and also shows superior performance in terms of MSE to conventional, habitual sleep diaries in this large cohort. Furthermore, the comparable results for the analysis carried out in the subset of the cohort with formally diagnosed sleep disorders point to the fact that our method may also be valuable when monitoring sleep in people suffering from these conditions. To the best of our knowledge, this is the first study that conducts these types of sensitivity analyses on a subset of disease subjects to show the validity of the proposed method in individuals who suffer from sleep disorders. Future work should carry more through validation on a larger population sample with sleep disorders where data has been collected in free-living conditions. This would allow additional insights into sleep disorders by leveraging the HR method’s ability to detect naps and daytime sleep.

We further examined the performance of the HR algorithm in the PhysioNet Apple Watch cohort that recorded concurrent Apple Watch data and a night of PSG. This study followed a similar experimental protocol to that of the MESA study, including the use of the BBVS night-only optimal parameter benchmark. In this cohort, the HR algorithm yielded an MSE of 0.07 and a time-deviation of -29.07 minutes when compared to gold-standard measures of sleep through PSG. These results showcase the potential of the method in commercial-grade wearable devices that obtain HR through photoplethysmography (PPG). Our approach is likely to translate well to other commercial, PPG based devices as they have shown to be reliable at detecting resting and sleeping heart rate, which are critical for our method and being less reliable for higher heart rates due to artifacts associated to vigorous activity [42]. We also examined the angle change approach in this cohort, with this method performing less well than it did in BBVS, yielding an MSE of 0.12 and a total sleep time deviation of 44.39 minutes.

Finally, we tested our method in the MMASH cohort, where free-living HR, movement and sleep diary data was available for entire days. As additional validation, we split this data into full-day and night-only subsets to replicate the BBVS analysis. Using BBVS-derived benchmark parameters for these 2 data subsets, the MSEs against sleep diaries were 0.11 for the full-day and 0.09 for the night-only analyses. For the full-day data, the total sleep time deviation of 17.64. The angle change approach resulted in an MSE of 0.10 and total time deviation of -55.86.

The results validate the versatility of the HR algorithm across multiple datasets. Prederived optimal paratemers can be used quickly on other cohorts, with the caveat of using the appropriate full-day and night-only values, due to the sleep time prior built into the *Q* parameter. If the lowest possible MSE value is desired, then the analysis can be further optimized with a cohort-specific parameter search.

One important limitation of the BBVS and MMASH studies is that they did not include PSG-derived ground truth sleep annotations. Although an ideal experimental protocol would have multiple days of PSG and free-living wearable sensor data, detailed sleep diaries allowed us to evaluate the algorithm across more than one or two nights, showcasing the strength of our method in discerning both inter- and intra-individual variability. Similarly, the accelerometers included in these studies offer an important perspective on accelerometry-based angular postural changes and how they compare to our proposed approach. Moreover, in ideal circumstances, HR for the full day would have been available in both the MESA and PhysioNet cohorts, optimizing the results of our approach by having exposure to non-sedentary wake behaviors. However, the results in these two datasets showcase the validity of our approach even under constrained laboratory conditions.

Future work should explore the robustness of the HR-based algorithm in cohorts such as inpatients. As the algorithm relies on HR signals already monitored continuously for other medical purposes, no additional accelerometer sensor would be required. Accurately labeling sleep in inpatients is challenging due to other factors that influence the HR ECDF, such as limited mobility, fever, medication, physiological and psychological stress, drug and alcohol use and cardiovascular conditions. However, objectively monitoring sleep without additional obtrusion could help improve sleep quality during hospital stays, which is a challenge for most patients [43], and hence promote both healing and patient satisfaction. Moreover, optimization of the angle change approach should be explored such that it can be used more reliably in the absence of HR sensor data, in this investigation we limited our evaluation of this method to the original parameters reported [27]. Parameter optimization could yield more generalizable and stronger outcomes for this approach. Finally, our method could be used in collaboration with some of the well-established activity-based approaches where multimodal settings are present. For instance, using conditional programming traditional methods could complement our approach in the detection of awakenings and assist in the derivation of conventional and novel sleep metrics.

Overall, our work highlights the potential of HR to detect the sleeping window not only in research and clinical contexts, but also in ecologically valid free-living conditions, enabling the objective monitoring of sleep in large-scale populations without PSG labels or sleep diary guidance. While there is value in algorithms or models that can be improved with self-report sleep data, the low effort involved in collecting and analysing objectively inferred sleep data through our method, coupled with low exclusion rate due to technical issues, missed diary entries or dropout would likely result in larger and more diverse study cohorts, as well as facilitating long-term objective data collection. For instance, few studies have been able to properly test the longitudinal, and likely synergistic, association between sleep quality and disease. Where this has taken place, sleep data is often collected through questionnaires [44] or with short, arbitrary follow-up periods. These studies could have missed long-term trends that significantly influence health status over months or years.

In sum, our proposed method was shown to be a valuable device-agnostic tool to infer sleep in both free-living and laboratory conditions without the need for sleep diaries. As highlighted by Depner and colleagues [45], our analysis and evaluation will help enable the translation of findings from laboratory-based sleep studies into large-scale cohort studies and clinical trials, by providing an objective, device agnostic method to monitor sleep without the need for sleep diaries.

## 4. Methods

### 4.1. Data Sources and Processing

In this study we used four different data sources with a variety of devices and populations to showcase the performance of our proposed method. Table 8 summarizes the types of wearable devices and ground truth used in each one of the studies.

**Table 8:** Summary of population size and devices used in the different datasets.

| Study | # Participants | Sensor type | Wearable device make | PSG | Sleep Diary |
| --- | --- | --- | --- | --- | --- |
| *Biobank Validation Study* | 158 | Triaxial accelerometer (3)<br>Wearable ECG | AX3, Axivity (Newcastle,UK)<br>Actiheart, CamNtech (Cambridge,UK) |  | ✓ |
| *Multi-Ethnic Study of Atherosclerosis* | 1154 | Actigraphy monitor<br>ECG | Actiwatch Spectrum, Philips Respironics (PA,USA) | ✓ | ✓ |
| *PhysioNet Apple Watch* | 22 | Triaxial accelerometer<br>Heart rate sensor (PPG) | Apple Watch (Series 2,3), Apple (CA, USA) | ✓ |  |
| *Multilevel Monitoring of Activity and Sleep in Healthy people* | 20 | Triaxial accelerometer<br>Heart rate sensor | ActiGraph wGT3X-BT, ActiGraph LLC (FL,USA)<br>Polar H7, Polar Electro Inc (NY,USA) |  | ✓ |

#### 4.1.1. The UK Biobank Validation Study (BBVS)

Participants of the BBVS study were recruited from the Fenland study [46]. In brief, 193 participants were recruited between the ages of 40 and 70, with a BMI between 20 and 50kg *·* m*^−^*^2^. Recruitment aimed to balance age, sex, and BMI distributions. Participants were invited to attend an assessment centre on two separate occasions, separated by a free-living period of 9 to 14 days during which they wore three waveform triaxial accelerometers (dominant and non-dominant wrists and thigh) as well as a combined movement and heart rate sensor. During the free-living period, participants were asked to keep a detailed log of their sleep, by recording the time they fell asleep and woke up on a daily basis. Ethical approval for the study was obtained from Cambridge University Human Biology Research Ethics Committee (Ref: HBREC/2015.16). All participants provided written informed consent. Full details of the BBVS study are described elsewhere [47].

Participants were fitted with a combined heart rate and movement sensor (Actiheart, CamNtech, Cambridgeshire, UK), measuring heart rate and uniaxial acceleration of the trunk every 15 seconds [48]. In addition, participants were fitted with three waterproof triaxial accelerometers (AX3, Axivity, Newcastle upon Tyne, UK); one device was attached to each wrist with a standard wristband, and one to the anterior midline of the right thigh using a medical-grade adhesive dressing. These devices were set up to record raw, triaxial acceleration at 100Hz with a dynamic range of *±*8*g*. BBVS participants were asked to wear all four devices continuously for the following 8 days and nights while continuing with their usual activities. In addition, they were asked to complete a diary of their sleep onset and wake times daily. This ensured that any small changes in onset and offset of sleep were captured during the recording period.

Following the download of the devices, the combined sensor heart rate data was cleaned and non-wear periods identified by the combination of non-physiological heart rate and prolonged periods of no movement [49]. All signals from the triaxial accelerometers were re-sampled to a uniform 100Hz signal by linear interpolation, and then calibrated to local gravity using a well-established technique [50, 51]. Periods of non-wear were classified on the basis of windows comprising an hour or more wherein the device was inferred to be completely stationary, where stationary is defined as standard deviation in each axis not exceeding the approximate baseline noise of the device itself (13*·*milli-*g*). All non-wear periods were removed from the analysis. Additionally, pitch, roll and z-angles for all three accelerometry devices were calculated enabling angular postural assessments and direct comparisons to previously established approaches which only rely on acceleration data [27, 41]. The residual acceleration signal can be interpreted as a measurement of the rotated gravitational field vector which can then be used to determine the accelerometer’s orientation angles (the conventional pitch and roll and z-angle, defined as the dorsal-ventral direction [27, 41]). Angles for each device were derived according to these formulae:

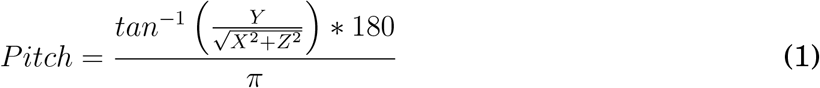

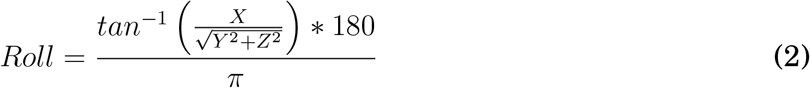

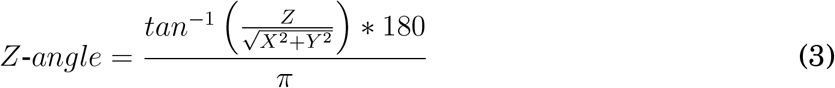

The accelerometry and heart rate signals were summarized to a common time resolution of one observation per 30 seconds and the time-series were aligned. Participants were excluded from the final analysis if they had less than 72 hours of concurrent wear data (three full days of recording from all four devices). Participants with less than 3 nights of concurrent wear and diary data were excluded from the final analysis. After these pre-processing steps, the resulting analytical sample was of 158 participants. Three of these participants were on cardioreactive medication and two were taking betablockers.

#### 4.1.2. Multi-Ethnic Study of Atherosclerosis (MESA)

The Multi-Ethnic Study of Atherosclerosis (MESA) is a multi-site prospective study that includes 6,814 men and women who identify as White, Black/African American, Hispanic, or Chinese, and are between the ages of 45-84 [52, 53]. Participants in this study were followed prospectively to evaluate risk factors for cardiovascular disease. 2,237 MESA participants are enrolled in a sleep exam (MESA Sleep Ancillary Study [54]), which includes seven days of wrist-worn actigraphy, one full overnight unattended polysomnography (wrist-worn actigraphy collected concurrently), and a sleep questionnaire. MESA participants who reported regular nighttime use of nocturnal oxygen or positive airway pressure devices were excluded from participation.

The MESA Sleep Study was conducted using a Compumedics Somte System for PSG, which includes the ECG signals here used to derive HR and HRV and their associated features, alongside an Actiwatch Spectrum from Philips Respironics (Pennsylvania, USA) to record actigraphy data. This device captures measurements of movement defined as “activity counts”^1^ and aggregates them into 30 second epochs. The Actiwatch was securely fastened to participant’s non-dominant wrist. These actigraphy signals and their associated features can be derived in most research-grade wearable devices. The sensors for the Compumedics PSG comprised: cortical EEG, bilateral EOG, chin EMG, abdominal and thoracic respiratory inductance plethysmography, airflow, ECG, leg movement sensor and finger pulse oximetry. These sensors collected three types of signals: bioelectrical potentials (EEG, EOG, EMG, ECG), waveforms received from transducers (thermistors on the airflow devices, inductance respiratory bands, piezo leg sensors and position sensors from the leg device) and auxiliary devices (oximetry measures of oxyhemoglobin saturation and nasal pressure records). Full details of the setup, protocol and sampling rates are available elsewhere^2,3^. All participants included in our study had at least one full night of PSG recording with concurrent actigraphy and ECG. All nocturnal recordings were transmitted to a centralized reading center at the Brigham and Women’s Hospital (Boston, MA, USA) and data was scored by trained technicians using AASM guidelines.

For this study, we synchronized PSG, ECG and actigraphy records into 30-second sleep epochs for a subset of 1,743 out of the 2,237 participants included in the original study. A total of 494 participants were excluded on the basis of: (1) lack of concurrent PSG, ECG and actigraphy data; (2) lack of sufficient quality standard data (*<*3h of usable data from the concurrent three sensing methods); or (3) lack of data integrity or misalignment of data, removing the resulting actigraphy outlier epochs based on human expert annotations. These outliers resulted from either non-wearing periods or equipment failure periods. For actigraphy epochs labeled as outliers, their corresponding HR/HRV epochs were also removed [55]. Further, given that some participants records comprised almost no PSG-labelled wake, which is unrealistic for free-living recordings and far removed from the general 24-hour HR quantile assumption of the algorithm, we only included participants who had at least 30 minutes of wake time prior to sleep onset and a maximum of 240 minutes after sleep offset, resulting in a total of 1,154 participants.

To obtain HR information, we used the QRS complexes (R-points) detected using Compumedics Somte (Abbotsford, VIC, Australia) software Version 2.10 (Builds 99 to 101). The R-points were classified as normal sinus, supraventricular premature complex or ventricular premature complex. For the data cleaning, filtering and noise removal, we used the Python package HRV-analysis^4^. First, RR interval outlier data was filtered using a threshold method, with a range between 300 to 2000 ms, based on the approach previously described by Tanaka et al. [56], then ectopic beats were removed by through the methods described in Malik et al. [57]. After this step was completed, we linearly interpolated the removed R-points and we grouped the RR intervals into 30 seconds epochs.

All data used from the MESA Sleep Ancillary study used in this work is publicly available from the National Sleep Research Resource repository^5^. Institutional Review Board approval was obtained at each MESA study site (Wake Forest University School of Medicine, Northwestern University, University of Minnesota, Columbia University, University of California Los Angeles and the Johns Hopkins University). All participants provided written informed consent.

A number of common sleep disorders were identified and logged for the MESA sleep study, representing numbers that are close to their real prevalence in similar populations. A breakdown of those diseases is presented in Supplementary Table S1.

#### 4.1.3. PhysioNet Apple Watch Polysomnography Study

Data for this study was collected at the University of Michigan between 2017 and 2019. The study consisted of 39 healthy subjects with no prior diagnosis of sleep-related breathing disorders, parasomnias, restless leg syndrome, central disorders of hypersomnolence, peripheral vascular disease, cardiovascular disease, vision impairments not correctable by glasses or contact lenses or other disorders that could cause neurological or psychiatric impairment. The study also excluded on the basis of shift work and recent transmeridian travel. Furthermore, participants were ruled out on the basis of excessive daytime sleepiness according to the Epworth Sleepiness Scale, and after the PSG visit, participants which showed symptoms of either obstructive sleep apnoea or REM sleep behaviours were also excluded. A total of 31 subjects met the required criteria. Data for the study can be obtained through Physionet [58] and a detailed description of this dataset is available elsewhere [31].

Participants in this study wore an Apple Watch to collect their activity patterns for 7 to 14 days before spending one night in a sleep lab. During the final night, participants underwent a PSG study while wearing the Apple Watch device (which collected HR and triaxial acceleration). The study was approved by the University of Michigan Review Board and all participants provided written informed consent.

For the PhysioNet Apple Watch study, Apple Watch raw triaxial acceleration data (x, y, z axis measured in *g*) at a 50Hz resolution was converted into angular postural based metrics like the ones described on BBVS. The Apple Watch measures HR in beats per minute, sampling every several seconds through its PPG sensor. For our analysis, we down-sampled HR by taking the mean of all samples within 15-second windows. For the PhysioNet Apple Watch study, the laboratory technicians started a “recording” period for the watch before the PSG recording started. For our final analysis, we only included participants whose sleep onset and offset were greater than 10 minutes from the start and end of the recording period, respectively. Through this process we intended to introduce a more realistic setting for our model. Details on the laboratory PSG settings can be found elsewhere [31]. The final cohort consisted of 22 participants.

#### 4.1.4. The Multilevel Monitoring of Activity and Sleep in Healthy people (MMASH)

Data for the MMASH study was collected by BioBeats in collaboration with researchers from the University of Pisa and was obtained through Physionet [58, 59]. The study collected data from 22 healthy young adult male participants comprising continuous heart rate and triaxial accelerometry monitoring as well as a variety of questionnaires to assess their physical activity, psychological and sleep characteristics as well as a detailed sleep diary. Participants also recorded their perceived mood (Positive and negative Affect Schedule-PANAS), Daily Stress Inventory (DSI) during the free-living protocol and completed a Morningness-Eveningness Questionnaire (MEQ), State-Trait Anxiety Inventory (STAI-Y), Pittsburgh Sleep Quality Questionnaire Index (PSQI) and Behavioural avoidance/inhibition (BIS/BAS) during their clinic visit. Further, anthropomorphic characteristics were recorded. All data was processed and recorded by sport and health scientists with the objective of assessing psychophysiological response to stress stimuli and sleep.

The 22 MMASH participants were fitted with two devices for continuous recording during 2 days: a heart rate monitor (Polar H7, Polar Electro Inc., Bethpage, NY, USA) which recorded beat-to-beat intervals and was used to obtain HR data and a triaxial accelerometer (ActiGraph wGT3X-BT - ActiGraph LLC, Pensacola, FL, USA) was worn on the wrist. Participants were asked to wear the devices continuously during the duration of the protocol and to complete a diary of their sleep onset and wake up times during the recording period. For MMASH we followed the same pre-processing, data quality and noise removal protocols that we described in BBVS for both the triaxial accelerometry signal and the HR signal. Two participants were removed from analysis on the basis of missing diary entries.

All participants provided written informed consent. Information was provided to them regarding the research protocol in accordance with General Data Protection Regulation: Regulation - EU 2016/679 of the European Parliament and of the Council 27/04/2016. Further, all experiments conducted were in accordance with the Helsinki Declaration as revised in 2013, the study was approved by the Ethical Committee of the University of Pisa (#0077455/2018).

### 4.2. Algorithm to estimate the sleep window using heart rate

Several challenges must be accounted for when developing a method for the detection of sleep in free-living conditions. First and foremost, most methods derived for sleep-wake classification using wearable devices have been derived on and for use during the night period [17, 19, 20, 23, 31]. These approaches were mostly conducted in small studies using concurrent PSG and as such, their application during the full day period greatly compromises the quality of the results. They also tend to be optimized in small, non-diverse populations, comprising their generalizability to other cohorts. Moreover, they tend to be device and make specific, often requiring conversions into arbitrary activity intensity measures or counts. Finally, most algorithms that can be applied during the 24 hour period require sleep diaries or questionnaires for guidance, which are often biased and burdensome to obtain [60].

Here we introduce a simple approach to estimate sleep window leveraging the HR sensing capabilities that most modern wearables have. One of the major challenges presented by large cohort studies is inter-individual differences. For instance, individuals who are fitter, tend to have lower resting heart rates than those who are not as fit [61]. Hence, an approach that relies on HR signals should not follow a *one size fits all*, but rather adapt to each individuals’ own heart rate profiles. To account for these considerations, we use the empirical cumulative distribution function (ECDF) of each individual’s daily heart rate profile. This function, *F* (*x*), is the probability that for each individual their heart rate takes a value *x* such that:

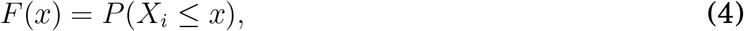

for every sequence *i* = 1*, …, n*. Namely, *F* (*x*) is the probability of the event *{X_i_ ≤ x}*. In this case, *x* is a threshold heart rate value (in beats per minute). To estimate the probability of a given event, we turn to the ratio of such an event given an individual’s daily sample of heart rates. This results in:

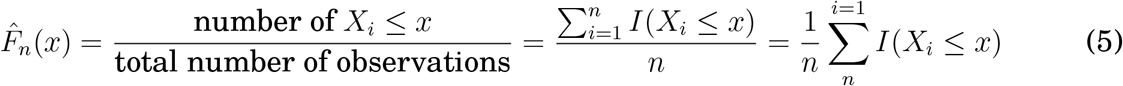

as the estimator of *F* (*x*), that is the ratio of HR less than *x*, where *I*() is the *indicator function*.

Thus, for every *x*, we can use such quantity as an estimator, so the estimator of the cumulative distribution function, *F* (*x*) is *F*^^^(*x*), which is referred to as the *empirical cumulative distribution function*.

By using the HR cumulative distribution function for each participant and each day of recording, our method accounts for inter- and intra-individual variation. It can adjust to different levels of fitness which often result in different resting HR during sleep [61]. Further, an elevated resting heart rate (RHR) accompanied by a fever is a well-known response to infection [62], alcohol consumption [63], stress [64] and can even be used to monitor influenza-like illness [65], something that our approach would account for. The method contains no in-built assumption of absolute time for the sleep window, and can therefore be used in night shift-workers and non-monophasic sleepers (those whose have more than one principal sleep windows in a 24-hour period) where the circadian HR rhythm is shifted so that most of the lower HR values still occur during sleep independent of the absolute time window when their sleep takes place. An example of our method applied to a shift worker can be observed in Supplementary Figure S1.

The first step of our heart rate sleep algorithm involves pre-processing the time series by assigning binary wake/sleep labels whenever the participant’s heart rate dips above/below a specific **quantile threshold (***Q*). The threshold value is calculated from the ECDF over 24-hour windows arbitrarily starting at 15:00 each day. Figure 4 showcases this cutoff for the full BBVS population based on two intervals (full day and from 21:00 to 11:00, a conventional night).

Wake/Sleep labels are then smoothed with a 5-minute rolling median and the length of their sequences is calculated. Sequences of sleep labels that are longer than a **minimum length (***L*) are extracted and merged with other sleep sequences if their **gap length (***G*) is smaller than a pre-defined value. We study the behavior of the parameters *Q*, *L* and *G* for each dataset with the goal of finding the best possible combination.

To be eligible as part of the final sleep window, the sleep sequence must not be preceded by more than 90 minutes of wake in the previous 4 hours of recording. The limits of the merged sleep sequences then guide a search (in a window starting 240 minutes before and 60 minutes after) for epochs with high HR volatility. This HR volatility threshold is defined as a rolling 10-minute standard deviation of the HR signal of 6 beats per minute. Defining the final sleep window limits as the last, and first high volatility epochs for sleep onset and offset, respectively, is meant to increase the algorithm’s sensitivity at discriminating sedentary time just before or after sleep (e.g., reading in bed) from the sleep window itself.

Finally, the algorithm also labels naps and awakenings, but these were not used in the analysis of the present datasets. Naps are the initial sleep sequences that lie outside a buffer 180-minute window either side of the main sleep window. For awakenings, the algorithm labels all the epochs when the HR rises above a quantile threshold *AV* extracted from the daytime (8am - 10pm) HR ECDF. From these only the sequences longer than 5 minutes are kept and then the sequences separated by less than 5 minutes of sleep are merged and then labeled as the final awakenings.

Pseudocode for the approach is provided in the Supplementary Material 1. A visual overview of the algorithm is provided in Figure 5 and Figure 6 showcases the application of the algorithm to a random participant trace.

### 4.3. Evaluation of the proposed approach

We used the four previously described cohorts to evaluate our method against gold-standard measures of sleep using PSG (MESA, Apple Watch PhysioNet) and detailed silver-standard measures through sleep diaries, as opposed to habitual sleep diaries which could be subject to recall bias (BBVS, MMASH). Although an ideal experimental protocol would have multiple days of PSG and free-living wearable sensor data, detailed sleep diaries allowed us to evaluate the algorithm across more than one or two nights, showcasing the strength of our method at discerning both inter- and intra-individual variability.

We performed epoch by epoch evaluation on all four cohorts and derived comparisons regarding the performance of our method with regards to total sleep time (TST), sleep onset and sleep offset time.

#### 4.3.1. Evaluation metrics

The following performance metrics were used to evaluate against the ground truth in each study: differences in onset/offset/total sleep block duration (minutes), mean square error (MSE) and Cohen’s *κ*. We evaluated our algorithm systematically for individual HR CDF quantiles *Q ∈* [0.10, 0.95] with step size 0.025, window lengths *L ∈* [10, 120] minutes with step size of 5 minutes, and gap between blocks *G ∈* [30, 420] minutes with step size of 30 minutes, optimizing for MSE.

We defined MSE as:

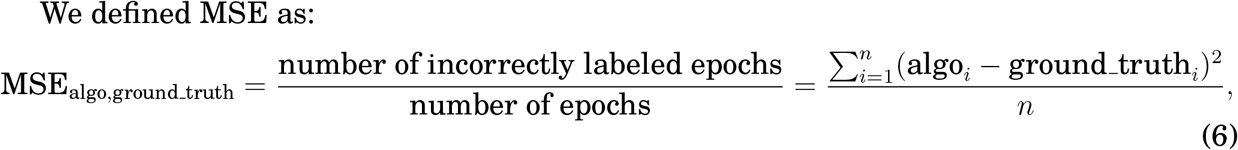

where *algo* and *ground_truth* are the binary labels for an epoch (1 for sleep, 0 for wake) out of *n* epochs in each subject’s time series. Epoch length is specified by the different study cohorts (1 minute in BBVS, 30 seconds in MESA and 15 seconds in both PhysioNet Apple Watch and 5 seconds in MMASH). Thus, if the sleep windows found by the HR algorithm match the ground truth labels exactly, *MSE* = 0. If the algorithm labels all epochs as wake, then MSE is the proportion of sleep in the time series according to ground truth, while if the algorithm and ground truth labels diverge entirely, MSE will be the sum of their sleep proportions out of the total time series. For all four cohorts we performed systematic parameter optimization for best MSE on the basis of quantile, window length and window merge values. We also computed Cohen’s kappa, which is used to determine the classifier agreement with ground truth (PSG or sleep diary), relative to chance [66]. Cohen’s kappa is calculated through (*p_o_ − p_e_*)*/*(1 *− p_e_*), where p*_o_* stands for the percentage of observed classifications with agreement, and p*_e_* is the percentage of classifications from hypothetical chance agreement. Finally, tests of statistical significance were conducted using a two-tailed t-test [67].

#### 4.3.2. Evaluation with sleep diary and angle change: BBVS

In the BBVS study, participants wore a variety of wearable devices and recorded the time they went to bed and woke up on a daily basis, providing detailed sleep diaries. As such, we conducted two types of evaluations on this cohort.

##### Evaluation with sleep diary

First, we compared the performance of our method against those sleep diaries. For our evaluation, we only included participants who had filled out those diaries and had more than three days of concurrent sensing and diary data. We evaluated our model against the diaries in terms of total sleep time, sleep onset and offset.

##### Evaluation with angle change algorithm

We assessed the performance of our approach versus an angular change algorithm inspired by previous work [27, 41]. The angular change approach started with calculating the pitch, roll and z-angle using triaxial acceleration for the device being evaluated. To isolate the gravitational acceleration for each axis, we applied a low-pass filter (0.2 Hertz) to each of the three axes (X, Y and Z) of every recording being evaluated.

Pitch, roll and z-angles were then calculated and the difference between successive epoch values was then smoothed using a 5 minute median rolling window. A threshold method (*<*10*^th^* percentile of values in that given day *·* 15) was applied to both columns, dividing the time series into initial sleep and wake blocks.

Of these blocks, only those larger than 30 minutes were kept. Blocks separated by less than 60 minutes were then merged and the largest block was deemed as the main sleep block within the day [27].

Two different angular change evaluations were performed, first, the intersection of the epochs when both pitch and roll calculations agreed on a sleep label created a voting system for a more reliable final sleep window. Alternatively, z-angle only measures were used to generate those sleep metrics as previously described [27]. No significant difference was found when comparing the performance of these two different approaches, so we only report the values obtained from the z-angle measures. All the previous steps were done separately for each limb (dominant and non-dominant wrists, and thigh) on which BBVS participants wore a device.

In BBVS, HR is recorded continuously across the 24-hr period. Thus, the threshold quantile is expected to be lower the longer the sampling interval for the ECDF given that sleep occupies a smaller proportion of the total interval being evaluated. To evaluate the effect of the chosen ECDF, we analyzed the optimal thresholds and their associated results to better understand how parameter choice may affect the performance of our approach.

#### 4.3.3. Evaluation with polysomnography and sleep diary: MESA

##### Evaluation with polysomnography

The recording time for PSG started when the subject’s setup was complete, yielding a period of sedentary wakefulness prior to sleep onset. While in an ideal scenario the participant would have been subject to ground truth recording also during the day, this is not a possibility given the nature of PSG. However, this limitation was addressed by evaluating PSG against sleep diary on the same dataset and evaluating our approach against both PSG and diary data. For this evaluation we compared the resulting sleep blocks from PSG, defined as epochs where the participant was in either NREM (N1, N2, N3) or REM sleep, to the sleeping window obtained through our HR algorithm.

Further, in MESA, we explored how our algorithm performed in healthy participants versus participants with sleep disorders. To do so, we first evaluated in the full cohort (n=1,154) and then on the subset of participants with (n=189, 16.4%) and without (n=965, 83.6%) any sleep disorders. The goal of this analysis was to caution and inform about potential limitations that our metho may have when evaluating in diseased participants.

##### Evaluation with sleep diary

PSG derived sleeping windows were compared to sleep diary records in the MESA cohort. This comparison allowed us to further understand the deviations of habitual self-reported sleep to objectively monitored, ground-truth through PSG. For the evaluation we use the same metrics as previously explored in the evaluation against PSG.

#### 4.3.4. Evaluation with polysomnography and angle change: PhysioNet Apple Watch Polysomnography Study

##### Evaluation with polysomnography

The PhysioNet Apple Watch study provided a unique opportunity to test our method in a commercial-grade wrist-worn wearable sensor that was concurrently worn during PSG. For this study, we used the same evaluation method explored in MESA, exploring our method based on the night-time concurrent recordings of wearable HR and PSG.

##### Evaluation with angle change algorithm

Given the multimodal nature of the study, we evaluated both the HR based algorithm and the angular change based algorithm on this population. For this evaluation we followed the same procedure as previously described on BBVS.

#### 4.3.5. Evaluation with sleep diary and angle change: MMASH

In the MMASH study, participants wore an HR strap and triaxial wrist accelerometer and recorded detailed sleep diaries including the time they fell asleep and woke up, which was filled on a daily basis. For this cohort, we also conducted two types of evaluation following the procedures used during the BBVS evaluation.

##### Evaluation with sleep diary

First, we compared the performance of our method against the sleep diaries of each participant. We evaluated our approach against the sleep diaries in terms of total sleep time, sleep onset and offset.

##### Evaluation with angle change algorithm

Similar to our second evaluation in BBVS, we also assessed the performance of our approach against the angular change approach previously described.

### 4.4. Statistical analyses

The results are represented as the mean *±* 95% confidence interval around the mean. The modified Bland–Altman technique was applied to verify the similarities between the different methods. Significant tests were conducted with a two-sided t-test. All statistical analyses were performed with Python 3.8 and SciPy 1.4.1.

## Data availability

Data for MESA, PhysioNet Apple Watch and MMASH are publicly available through the following resources: 1) MESA: https://sleepdata.org/datasets/mesa, 2) Physionet Apple Watch: https://physionet.org/content/sleep-accel/1.0.0/heart_rate/ and 3) MMASH: https://physionet.org/content/mmash/1.0.0/. BBVS data is available from the first author upon reasonable request.

## Code availability

The implementation of the HR sleep period method described in this paper is available in an open-source Python 3.8 package (https://github.com/HypnosPy/HypnosPy). This package also provides with a number of tools to analyze a variety of wearable devices and derive sleep, circadian rhythms and physical activity inferences.

## Data Availability

Data for MESA, PhysioNet Apple Watch and MMASH are publicly available through
the following resources: 1) MESA: https://sleepdata.org/datasets/mesa, 2) Physionet
Apple Watch: https://physionet.org/content/sleep-accel/1.0.0/heart_rate/ and 3)
MMASH: https://physionet.org/content/mmash/1.0.0/. BBVS data is available from
the first author upon reasonable request.

https://physionet.org/content/sleep-accel/1.0.0/heart_rate/

https://physionet.org/content/mmash/1.0.0/.

## Acknowledgements

For BBVS, we thank all the participants and the staff from the MRC Epidemiology Unit Functional Group Teams. In particular we would like to thank Lewis Griffiths and Stefanie Hollidge for their contribution to data collection and data preparation. We thank all the teams involved in collection and open access management of the MESA, PhysioNet and MMASH cohorts. These initiatives allow for greater transparency and advancement of wearable-based objective monitoring of physical activity and sleep. We would also like to thank Emma Clifton for her insightful comments on the implications of objective monitoring of sleep in the field of epidemiology.

Our work was supported by GlaxoSmithKline and EPSRC through an iCase fellowship (17100053), the Embiricos Trust Scholarship of Jesus College Cambridge, and EPSRC through Grant DTP (EP/N509620/1). The work of KW is supported by the NIHR Cambridge Biomedical Research Centre (IS-BRC-1215-20014). The icons used in some of the figures are licensed under Creative Commons by thenounproject.com.

## Author contributions

IPP, MP, DS, CM and JP conceptualised the study. IP, MP, and JP designed the analysis. IP, MP, DS and JP developed and implemented the models for sleep classification. JP and IPP conducted the statistical analysis, with advice from SB and CM. IPP led the writing of the manuscript while all authors contributed to the interpretation of results, and to the editing and revision of the manuscript. KW, NW and SB enabled BBVS access and helped contextualize the findings from a clinical medicine and epidemiological perspective.

---

**Figure S1:**
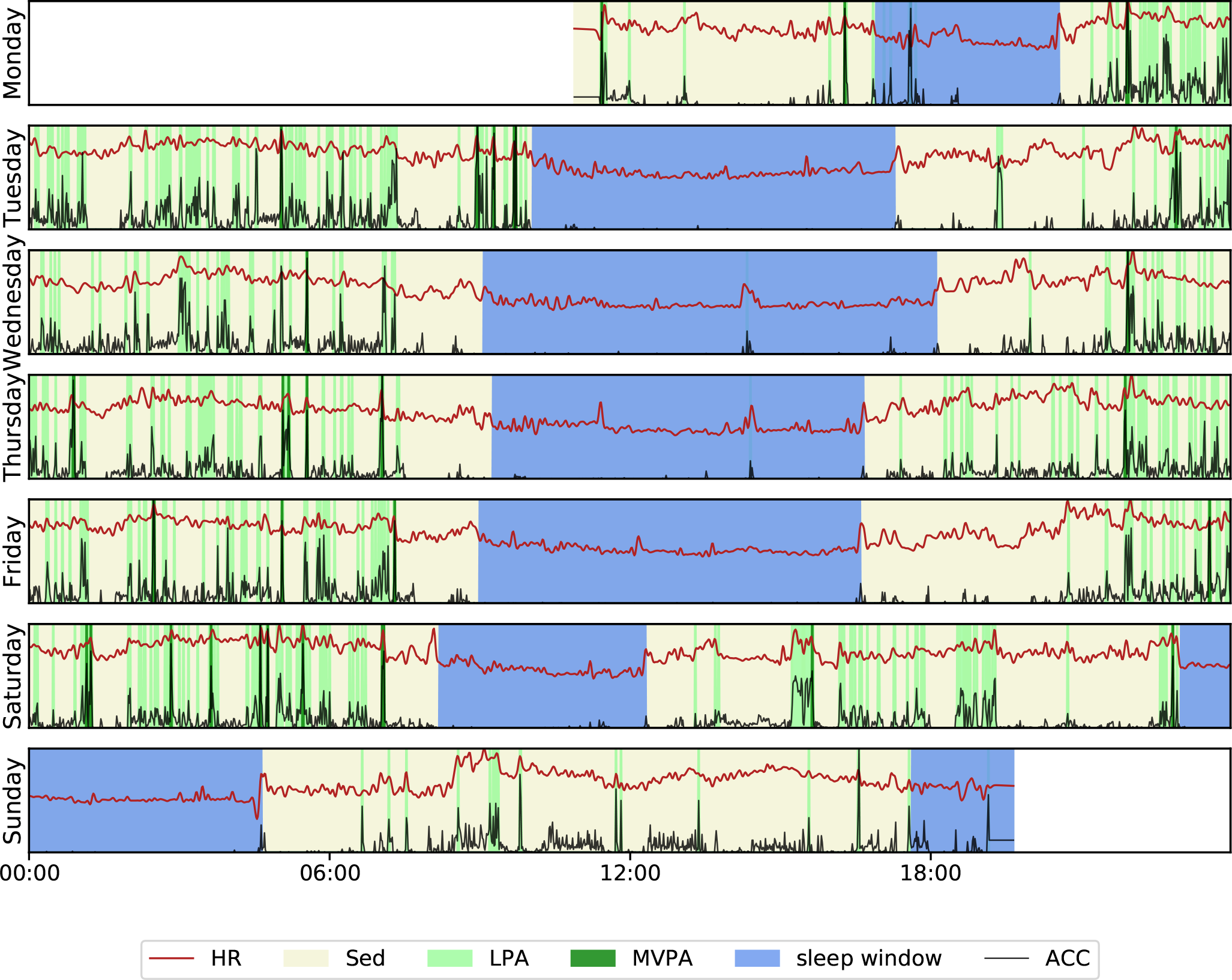
Applying the HR sleep algorithm on a shift worker. The free-living trace shows the subtle changes for day of the week picked up by the algorithm, with 2 sleep windows detected on Saturday, when they were not at work during the night. HR: Heart Rate; Sed: Sedentary; LPA: Light Physical Activity; ACC: Acceleration.

**Algorithm 1.**
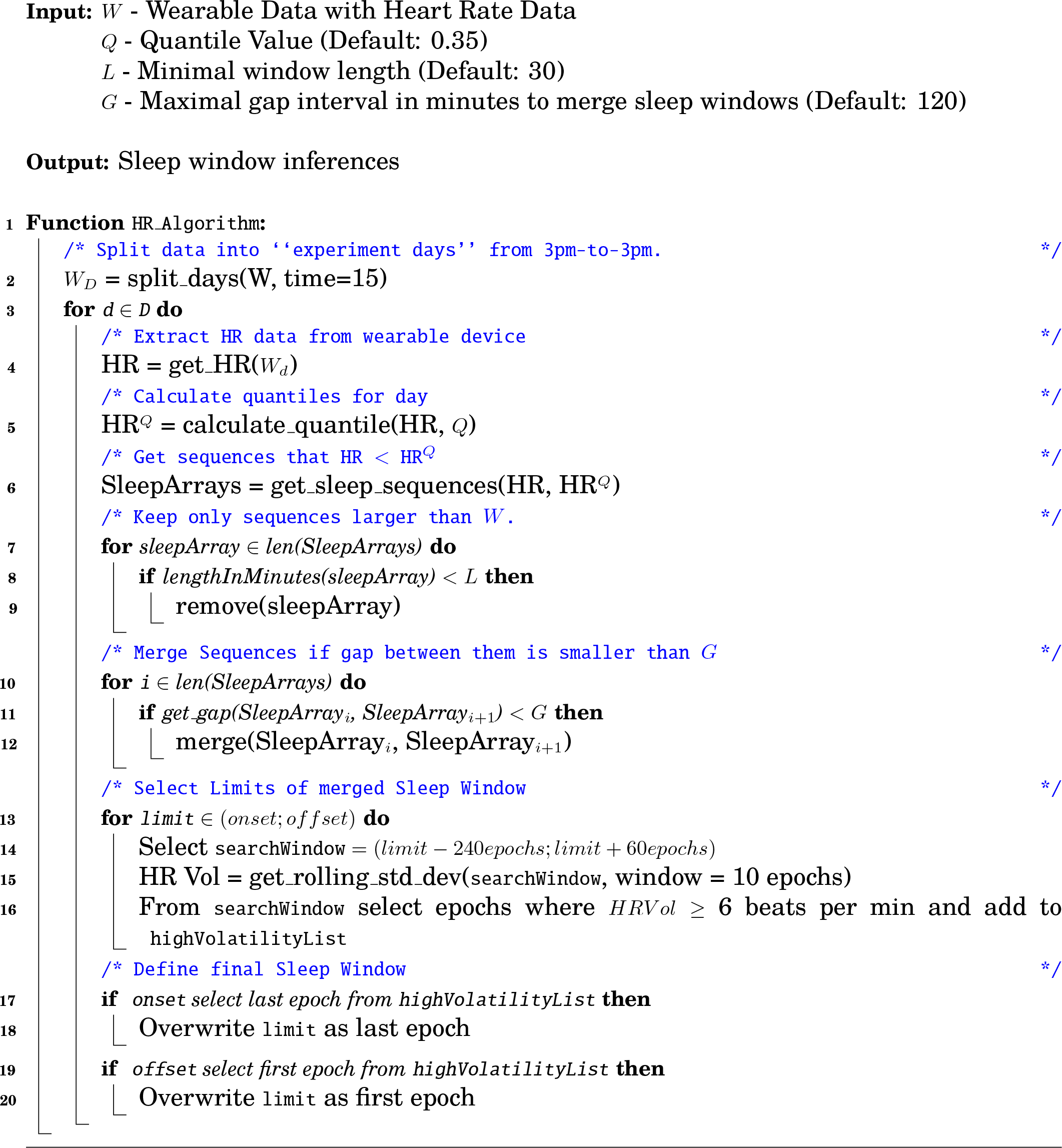
Method to estimate sleep periods based on Heart Rate.

**Table S1:** Prevalence of sleep disorders in the subset of MESA dataset used in this work (n=1,154) against the reported prevalence in the general population.

| Sleep Disease | Participants in Mesa (%) | Prevalence in the population |
| --- | --- | --- |
| Restless Legs Syndrome | 59 (5.11%) | 1.9 - 4.6% [68] |
| Insomnia | 67 (5.81%) | 6% [69] |
| Sleep Apnea | 95 (8.23%) | 3-7% [70] |
| Any of the above | 189 (16.38%) | - |

**Figure S2:**
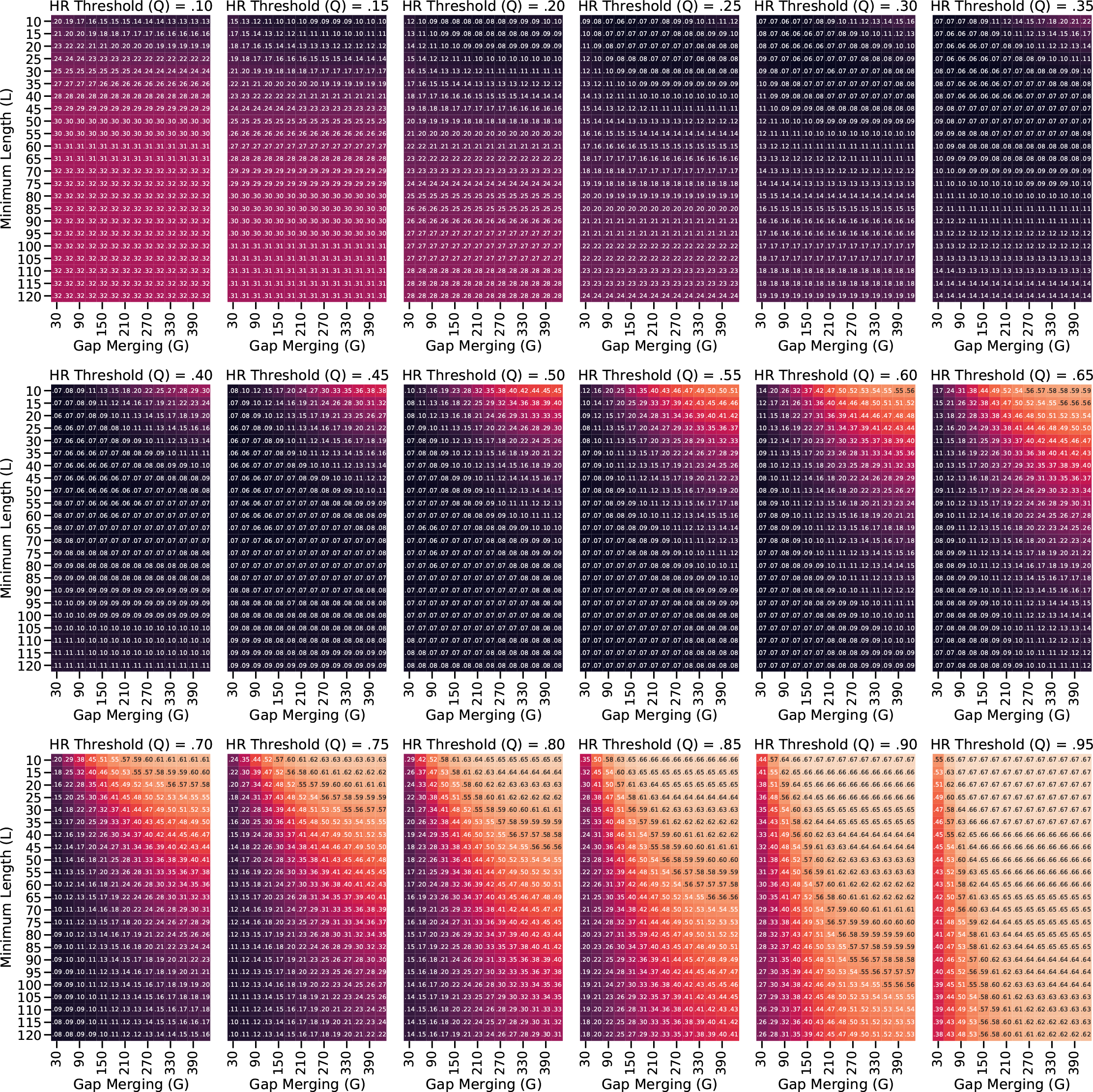
Details of the hyper-parameter search procedure for the full-day HR algorithm on the BBVS dataset.

**Figure S3:**
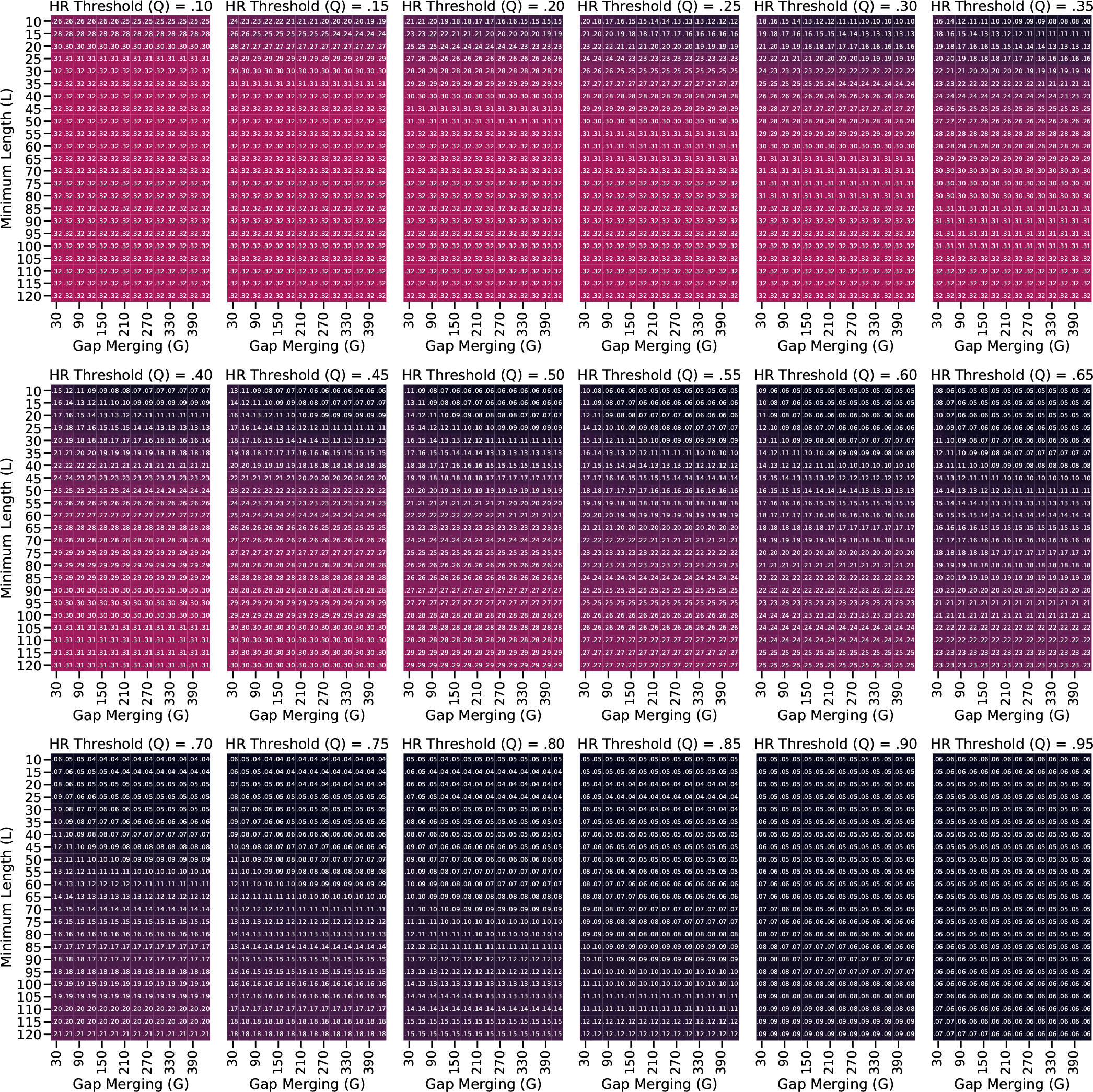
Details of the hyper-parameter search procedure for the night-only HR algorithm on the BBVS dataset.

**Figure S4:**
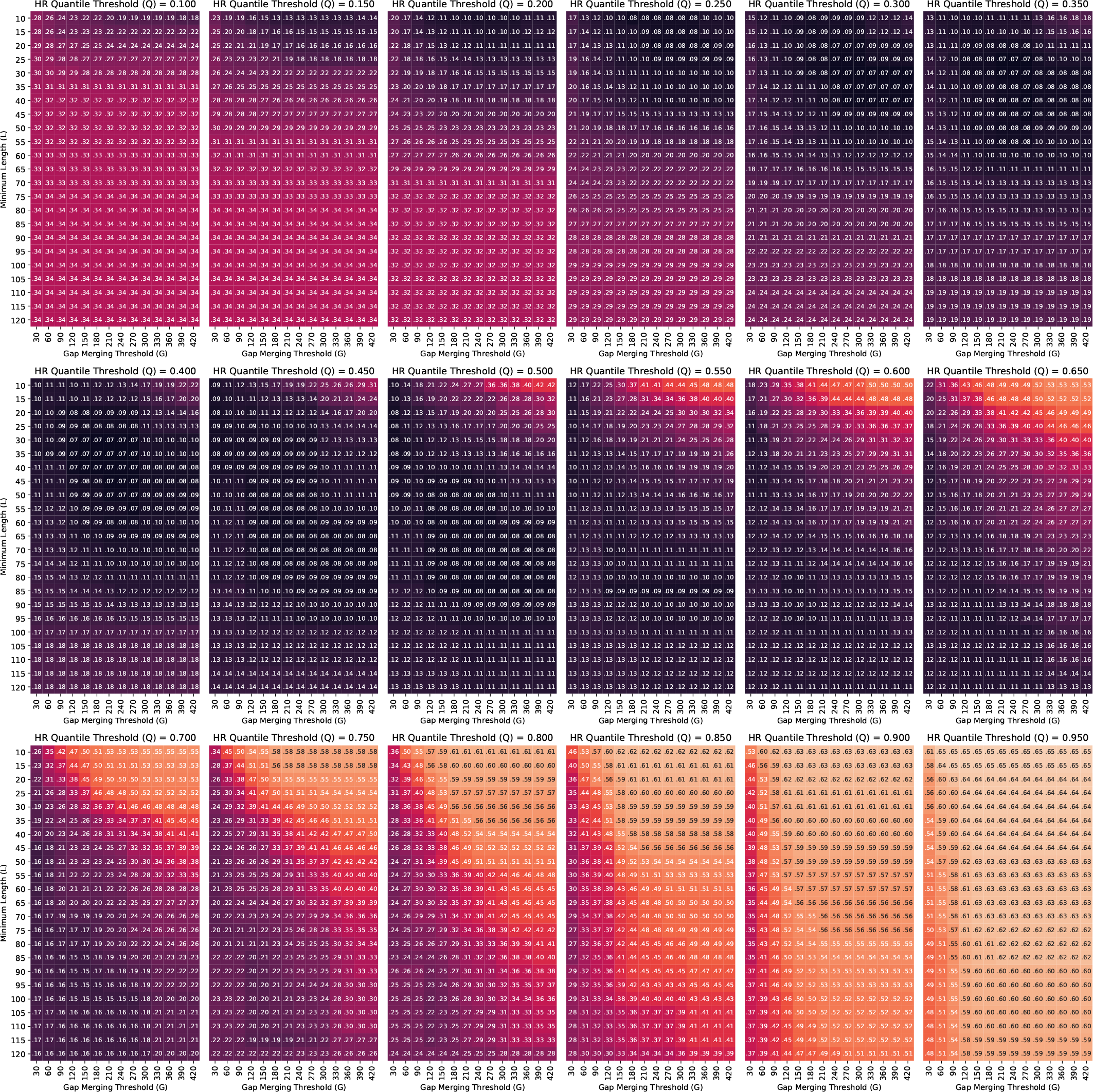
Details of the hyper-parameter search procedure for the full-day HR algorithm on the MMASH dataset.

## Footnotes

1 https://www.salusa.se/Filer/Produktinfo/Aktivitet/TheActiwatchUserManualV7.2.pdf

2 https://sleepdata.org/datasets/mesa/pages/equipment/montage-and-sampling-rate-information. md

3 https://sleepdata.org/datasets/mesa/files/documentation

4 https://pypi.org/project/hrv-analysis/

5 https://sleepdata.org/datasets/mesa

